# Synergizing Health Strategies: Exploring the Interplay of Treatment and Vaccination in an Age-Structured Malaria Model

**DOI:** 10.1101/2024.09.26.24314198

**Authors:** Mahmudul Bari Hridoy, Angela Peace

## Abstract

Malaria remains a persistent global challenge, particularly prevalent in tropical regions of Africa, Asia, and South America. According to the World Health Organization’s (WHO) World Malaria Report 2023, there were approximately 249 million reported malaria cases in 2022 across 85 endemic countries, resulting in over half a million deaths. Progress towards global malaria eradication through antimalarial drugs has been slow, with case numbers increasing since 2015. Dihydroartemisinin (DHA), artesunate, and artemether, derivatives of artemisinin, are crucial components of modern antimalarial treatment. However, resistance to these drugs and their partners in Artemisinin Combination Therapy (ACT) has emerged in Southeast Asia, Africa, and South America. In 2021, the WHO recommended widespread use of the RTS,S malaria vaccine among children in endemic regions. To address these challenges, we develop an extended SEIR age-structured model incorporating malaria vaccination for children, drug-sensitive and drug-resistant strains, and interactions between human hosts and mosquitoes. Our research focuses on evaluating how malaria vaccination coverage influences disease prevalence and transmission dynamics. We derive the basic, intervention, and invasion reproduction numbers for both strains and conduct sensitivity analysis to identify key parameters affecting infection prevalence. Our findings reveal that model outcomes are primarily influenced by scale factors that reduce transmission and natural recovery rates for the resistant strain, as well as by drug treatment and vaccination efficacies, and mosquito death rates. Numerical simulations indicate that while treatment reduces the malaria disease burden, it also increases the proportion of drug-resistant cases. Conversely, higher vaccination efficacy correlates with lower infection cases for both strains. These results suggest that a synergistic approach, involving both vaccination and treatment, could effectively decrease the overall proportion of the population that is infected.

## 1 Introduction

Malaria (derived from the Latin phrase *Malus aer*, meaning bad air) is a mosquito-borne disease primarily transmitted to humans through the bite of infected female Anopheles mosquitoes. Malaria is caused by a tiny protozoan parasite that falls under the Plasmodium species group, comprising various subspecies. Among the over 200 Plasmodium species, only five, namely *P. malariae, P. falciparum, P. vivax, P. ovale*, and *P. knowlesi*, are responsible for causing diseases in humans [2, 26]. Within the human body, these parasites undergo initial growth and replication within liver cells before moving on to infect red blood cells. Humans infected with malaria typically experience fever, fatigue, vomiting, and headaches, progressing through distinct phases known as the “hot,” “wet,” and “cold” stages. Malaria is a leading cause of illness and death in children, particularly in sub-Saharan Africa. Children under 5 are notably vulnerable due to their developing immune systems, with symptoms ranging from mild fever and cough to severe conditions like cerebral malaria, severe anemia, and respiratory distress [6, 8]. Timely identification and the correct course of treatment for malaria are essential to prevent illness and fatal consequences.

According to the World Health Organization’s (WHO) World Malaria Report 2023, approximately 249 million malaria cases were reported in 2022 across 85 endemic countries, marking an increase of 5 million cases compared to 2021. This surge in cases led to an estimated 608,000 deaths globally, corresponding to a mortality rate of 14.3 deaths per 100,000 people at risk [35]. 76% of these deaths were alarmingly among children under the age of five, meaning over a thousand young lives were lost every single day [31].

Dihydroartemisinin (DHA), artesunate, and artemether, which are derivatives of artemisinin, are now fundamental components of contemporary antimalarial treatment [12]. Artemisinin combination therapies (ACTs) and artemisinin derivatives are used to address mild cases of malaria. On the other hand, Artesunate (AS) is employed alongside Mefloquine in combination therapy to effectively manage moderate to severe cases of malaria. The derivatives of artemisinin are renowned for their effectiveness against P. falciparum malaria, which accounts for roughly 91% of global malaria cases and over 90% of malaria-related deaths in sub-Saharan Africa [20]. Unfortunately, resistance to Artemisinin derivatives and ACT partner drugs has notably emerged in Southeast Asia, Africa and South America [17, 22, 24]. Plasmodium species exhibit significant genetic adaptability to changes in their environment which gives them the capacity to rapidly develop resistance to treatments [26]. Progress in achieving global malaria eradication through antimalarial drugs has been slow, with a rise in case numbers since 2015. Therefore, it is crucial to preserve and enhance the efficacy of current antimalarial drugs due to the limited availability of new options.

In 2021, the WHO has issued a recommendation for the widespread use of the RTS,S malaria vaccine (brand name Mosquirix) among children living in malaria-endemic regions [1]. Although this vaccine showed some effectiveness in providing immunity to children under 5, it only offered partial protection [34]. The R21/Matrix-M™ vaccine, the latest advancement in malaria vaccination, improves upon the partial success of the RTS,S/AS01 vaccine observed in clinical trials. Demonstrating impressive efficacy, the R21/Matrix-M vaccine achieved a remarkable 77% effectiveness in Phase 2 clinical trials [23]. This cost-effective vaccine is administered in at least three doses to infants under two years old, with a booster (fourth dose) extending protection for additional years. As of October 2023, the vaccine has been administered to 2 million children residing in regions with moderate-to-high malaria transmission through the Malaria Vaccine Implementation Programme (MVIP) [36]. Additionally, at least 28 African countries intend to incorporate a WHO-recommended malaria vaccine into their national immunization programs. However, there is a noted decrease in vaccine effectiveness among older children [1]. Although the R21/Matrix-M™ Malaria Vaccine holds promise, it cannot address malaria alone. Effective malaria control requires a combination of interventions, including vector control, timely diagnosis and treatment, and, of course, the development of novel vaccines.

In the 1890s, Sir Ronald Ross used mathematical functions to study the link between mosquito populations and malaria incidence in humans [25]. Since then, numerous deterministic models have been developed to estimate the impact of vaccination on malaria transmission, incorporating host and parasite factors, their interactions, and environmental variables [7, 10, 11, 28, 29]. These models do not account for the maturation of the human age structure over time. On the other hand, age-structured models e.g. [9, 30] do not incorporate drug resistance into the model. More recently, Manore et al. [14] developed an age-structured ordinary differential equation model to assess how intermittent preventive treatment (IPT) impacts malaria-induced mortality and drug resistance in children. The model evaluates the effectiveness of IPT in reducing malaria deaths while considering the risk of promoting antimalarial drug resistance and the influence of drug half-lives on resistance spread. We extend this model to include vaccination for children and focus on antimalarial drug treatments, such as ACTs and artemisinin derivatives, to provide new insights.

In this manuscript, we extend prior analytical and computational research on mathematical modeling of malaria dynamics by developing an advanced SEIR age-structured model. This model incorporates malaria vaccination for children, both drug-sensitive and drug-resistant strains, and the interactions between human hosts and mosquitoes. Our research aims to evaluate the impact of malaria vaccination coverage on disease prevalence and transmission dynamics. The paper is organized as follows: Section 2 discusses the model formulation and outlines the variables. Section 3 provides an analysis of the model, including the derivation of the basic, intervention, and invasion reproduction numbers. Section 4 describes the parameters used and offers a sensitivity analysis of these parameters. Section 5 presents numerical explorations, and finally, Section 6 concludes the paper with remarks and suggestions for future research.

## 2 Model Formulation

We formulate a system of non-linear ordinary differential equations to represent an age-structured human population of Susceptible, Exposed, Infected, and Recovered individuals, considering vaccination and malaria treatment. The human population is classified into two age groups: individuals with naive immune systems, who are below 5 years old, and those with mature immune systems, who are 5 years old and above. The mosquito population is also modeled, where susceptible mosquitoes (*S*_*v*_) can become exposed and infected with the drug-sensitive malaria strain, transitioning from *E*_*s*_ to *M*_*s*_, or with the drug-resistant strain, transitioning from *E*_*r*_ to *M*_*r*_, respectively as seen in Fig: 1. The interaction between an infected mosquito and a susceptible human, with a transmission rate (*β*_*k*_), can result in the human contracting the sensitive parasite strain, denoted by the variable *I*, if the bite was from an *M*_*s*_-type mosquito. Conversely, if the bite was from an *M*_*r*_-type mosquito, it leads to the human being infected with the resistant parasite strain, identified by the variable *J*. Furthermore, we include a reduction factor, *κ* (0 *< κ <* 1), which scales down the transmission rates for individuals infected with the resistant parasite strain, whether they are mosquitoes (*κ*_*v*_) or humans (*κ*_*h*_). The variables and parameters values of the model are presented in the following Tables 1 and 2, respectively.

**Figure 1:**
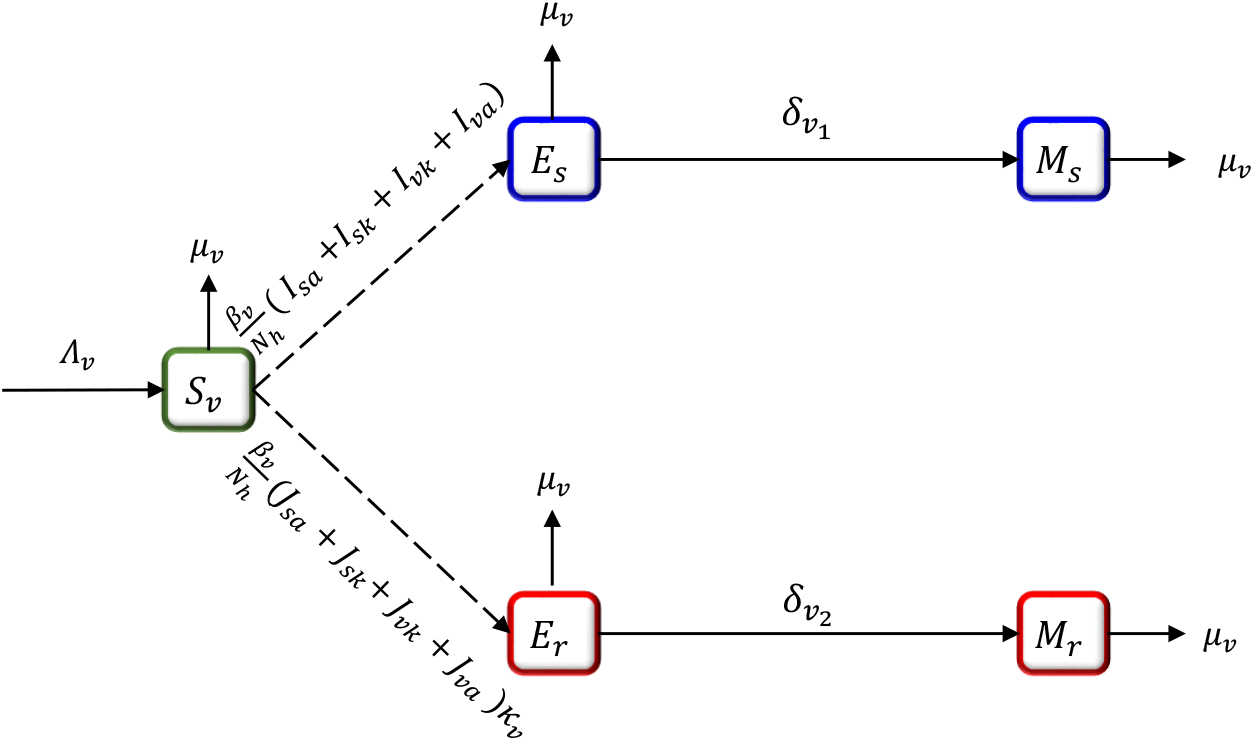
Transfer diagram for mosquito infection transmission dynamics.

**Table 1:** Description of the model state variables.

| Variable | Description |
| --- | --- |
| $S_k$ | Unvaccinated susceptible naive humans |
| $S_a$ | Unvaccinated susceptible mature humans |
| $S_v$ | Susceptible mosquitoes |
| $V_k$ | Vaccinated susceptible naive humans |
| $V_a$ | Vaccinated susceptible mature humans |
| $E_{sk}$ | Unvaccinated naive humans exposed to sensitive strain |
| $F_{sk}$ | Unvaccinated naive humans exposed to resistant strain |
| $E_{sa}$ | Unvaccinated mature humans exposed to sensitive strain |
| $F_{sa}$ | Unvaccinated mature humans exposed to resistant strain |
| $E_{vk}$ | Vaccinated naive humans exposed to sensitive strain |
| $F_{vk}$ | Vaccinated naive humans exposed to resistant strain |
| $E_{va}$ | Vaccinated mature humans exposed to sensitive strain |
| $F_{va}$ | Vaccinated mature humans exposed to resistant strain |
| $E_s$ | Mosquitoes biting humans exposed to sensitive strain |
| $E_r$ | Mosquitoes biting humans exposed to resistant strain |
| $I_{sk}$ | Unvaccinated naive humans infected with sensitive strain |
| $J_{sk}$ | Unvaccinated naive humans infected with resistant strain |
| $I_{sa}$ | Unvaccinated mature humans infected with sensitive strain |
| $J_{sa}$ | Unvaccinated mature humans infected with resistant strain |
| $I_{vk}$ | Vaccinated naive humans infected with sensitive strain |
| $J_{vk}$ | Vaccinated naive humans infected with resistant strain |
| $I_{va}$ | Vaccinated mature humans infected with sensitive strain |
| $J_{va}$ | Vaccinated mature humans infected with resistant strain |
| $M_s$ | Mosquitoes biting humans infected with sensitive strain |
| $M_r$ | Mosquitoes biting humans infected with resistant strain |
| $T_{sa}$ | Unvaccinated mature humans who have received treatment |
| $T_{va}$ | Vaccinated mature humans who have received treatment |
| $R_s$ | Recovered unvaccinated mature humans with temporary immunity |
| $R_v$ | Recovered vaccinated mature humans with temporary immunity |

**Table 2:** Model parameter description and values. NH: naive human, MH: Mature human, UV: Unvaccinated, V: Vaccinated, SS: Sensitive strain, RS: Resistant strain, * Assumed, ** Varied. Parameter for recovery rate (*γ*) was used for recovery rates for treated UV MH (*γ*_*sa*_), V MH (*γ*_*va*_), UV NH (*γ*_*s*_), and V NH (*γ*_*v*_). SC: Source.

|  | Parameter Description | Baseline value [Range] | SC |
| --- | --- | --- | --- |
| $N_h(0)$ | Initial human population | $5(10^6)$ | [21] |
| $N_v(0)$ | Initial mosquito population | $3 \cdot N_h(0)$ | [21] |
| $\Lambda_h$ | Recruitment rate into the susceptible NH | $3.55(10^3) [(2.24 - 5.08)10^3]$ | [21] |
| $\Lambda_v$ | Mosquito birth rate | $3 \cdot N_h/\mu_v$ | [14] |
| $\beta_k$ | Transmission rate of SS from mosquitoes to NH | 0.45 [0.18 – 0.9] | [14] |
| $\beta_a$ | Transmission rate of SS from mosquitoes to MH | 0.50 [0.18 – 0.9] | [14] |
| $\beta_v$ | Transmission rate of SS from humans to mosquitoes | 0.15 [0.03 – 0.2] | [14] |
| $\delta_{k_1} (\delta_{k_2})$ | Incubation rate for NH exposed to SS (RS) | 0.053 [0.033 – 0.143] | [4] |
| $\delta_{a_1} (\delta_{a_2})$ | Incubation rate for MH exposed to SS (RS) | 0.083 [0.033 – 0.143] | [4] |
| $\delta_{v_1} (\delta_{v_2})$ | Incubation rate for mosquitoes exposed to SS (RS) | 0.053 [0.029–0.33] | [5] |
| $\kappa_h$ | Reduction factor of human transmission rate by RS | 0.6 [0 – 1] | * |
| $\kappa_v$ | Reduction factor of mosquito transmission rate by RS | 0.6 [0 – 1] | * |
| $\xi_{I_{sa}}$ | Proportion of recovered UV MH from SS with immunity | 0.8 [0.1 – 1] | [21] |
| $\xi_{J_{sa}}$ | Proportion of recovered UV MH from RS with immunity | 0.8 [0.1 – 1] | [21] |
| $\xi_{I_{va}}$ | Proportion of recovered V MH from SS with immunity | 0.9 [0.1 – 1] | [21] |
| $\xi_{J_{va}}$ | Proportion of recovered V MH from RS with immunity | 0.9 [0.1 – 1] | [21] |
| $m$ | Maturation rate of NH | 0.000548 | [3] |
| $\mu_k$ | Natural death rate of NH | $5.94(10^{-4}) [(4.58 - 6.92)10^{-4}]$ | [14] |
| $\mu_h$ | Natural death rate of MH | $4.43(10^{-5}) [(4.25 - 4.79)10^{-5}]$ | [14] |
| $\mu_v$ | Natural death rate of mosquitoes | 0.07 [0.048 – 0.143] | [14] |
| $\nu$ | Vaccination rate of susceptible NH | 0.0085 | * |
| $P_{sk}(P_{vk})$ | Drug treatment efficacy of SS for UV (V) NH | 1 [0 – 1] | ** |
| $P_s(P_{vs})$ | Drug treatment efficacy of SS for UV (V) MH | 1 [0 – 1] | ** |
| $P_r(P_{vr})$ | Drug treatment efficacy of RS for UV (V) MH | $0.2 \cdot P_{vs}$ | ** |
| $\frac{1}{a}$ | Avg. time to clear sensitive strain | 5 [3 – 10] | [14] |
| $\omega$ | Rate of loss of natural immunity for UV MH | 0.003 [0.001 – 0.05] | [19] |
| $\omega_v$ | Rate of loss of natural immunity for V MH | 0.003 [0.001 – 0.05] | [19] |
| $\gamma$ | Recovery rate | 0.04 | [19] |
| $d_{sk} (d_{vk})$ | Disease-induced death rate for UV (V) NH | 0.0000101 | [14] |
| $d_{sa} (d_{va})$ | Disease-induced death rate for UV (V) MH | 0.00000109 | [14] |
| $\sigma_{sa} (\sigma_{va})$ | Natural recovery rate of UV (V) MH | 0.03[0.002 – 0.05] | [14] |
| $\phi$ | Scale factor reducing natural recovery rate for RS to SS | 0.5 | * |
| $\psi$ | Scale factor decreasing incubation period for VH | 1.1 | * |
| $\epsilon$ | Vaccination efficacy | 0.75 [0 – 1] | ** |

An unvaccinated susceptible naive human (*S*_*k*_) can either be exposed to the sensitive malaria strain and transition to (*E*_*sk*_), or be exposed to the resistant strain and transition to (*F*_*sk*_), see Figure 2 for model flow diagram. From (*E*_*sk*_), they can become infectious and transition to (*I*_*sk*_) for the sensitive strain, and from (*F*_*sk*_) to (*J*_*sk*_) for the resistant strain, with an incubation rate of *δ*_*k*_. Infected naive humans can naturally recover with a recovery rate (*γ*_*s*_) or through drug treatment with efficacy (*P*_*sk*_). For the resistant strain, the recovery occurs at a slower rate, with a scale factor (Φ) reducing the rate of natural clearance of resistant parasites relative to sensitive parasites. Unvaccinated naive humans can be vaccinated at a vaccination rate (*ν*) to transition to the vaccinated state (*V*_*k*_). We consider the vaccine to have a vaccination efficacy denoted by *ϵ*, which also creates a scaling factor, *ψ*, that increases the incubation rate from exposed to infectious for vaccinated humans. Similar to *S*_*k*_, a vaccinated susceptible naive human (*V*_*k*_) can either be exposed to the sensitive malaria strain and transition to (*E*_*vk*_), or be exposed to the resistant strain and transition to (*F*_*vk*_) and subsequently to (*I*_*vk*_) for the sensitive strain, or to (*J*_*vk*_) for the resistant strain. Infected vaccinated naive humans can naturally recover with a recovery rate (*γ*_*v*_) or through drug treatment with efficacy (*P*_*vk*_).

**Figure 2:**
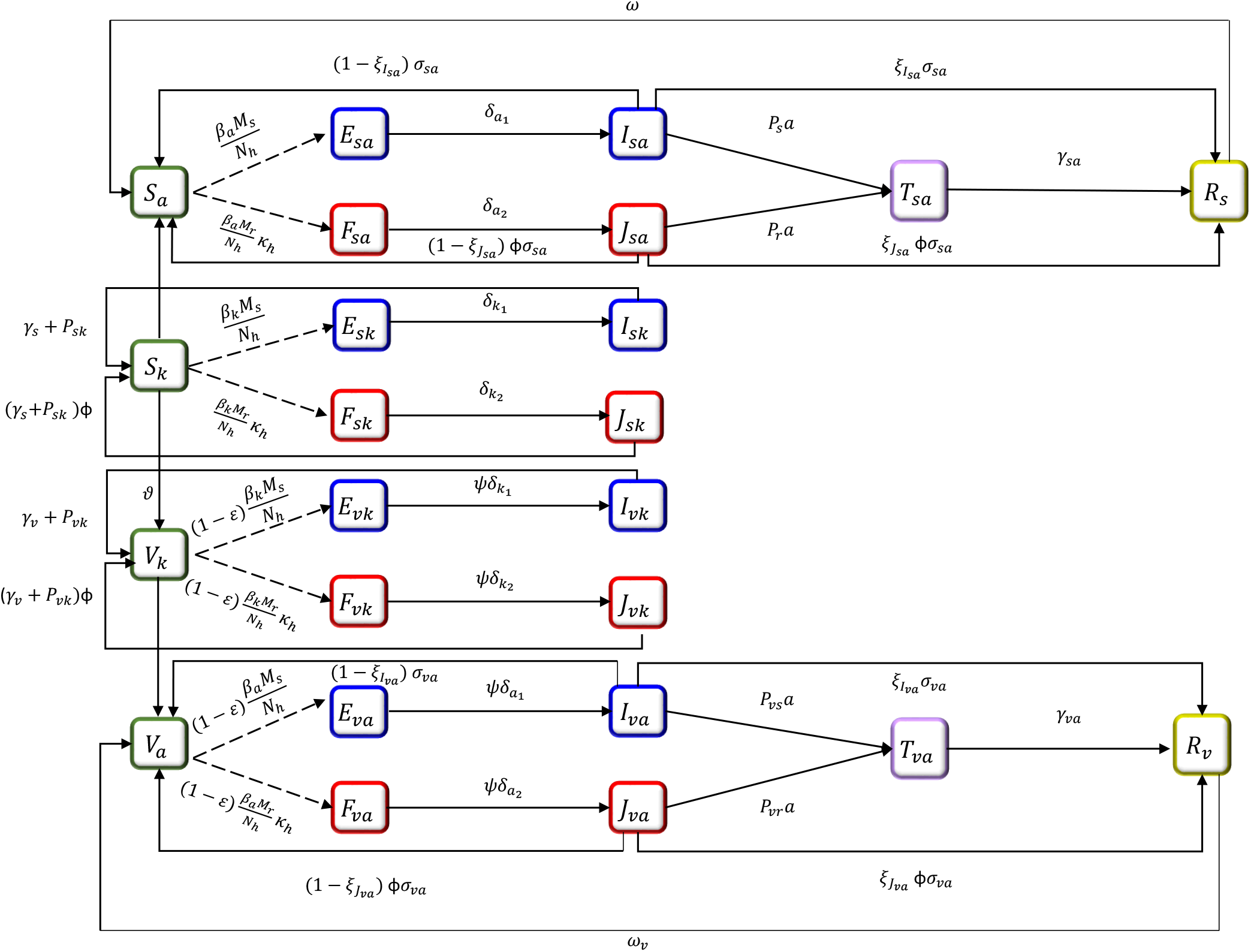
Transfer diagram for human infection transmission dynamics.

Each state for naive, unvaccinated, or vaccinated humans can transition to the corresponding mature class with a maturation rate (*m*), see Figure 3. Natural death rates are included as well: (*μ*_*k*_) for naive humans and (*μ*_*h*_) for mature humans, along with disease-induced death rates (*d*). For example, an unvaccinated, naive human infected with the sensitive strain (*I*_*sk*_), can experience natural death at rate (*μ*_*k*_), disease-induced death at rate (*d*_*sk*_), or they can mature to *I*_*sa*_, where they encounter a natural death rate (*μ*_*h*_) and disease-induced death rates (*d*_*sa*_). To simplify, only unvaccinated susceptible individuals can receive the vaccine, as it has been developed for children under the age of 5. Therefore, mature adults can only be vaccinated if they were vaccinated before the age of 5 and have since matured into the corresponding adult class.

**Figure 3:**
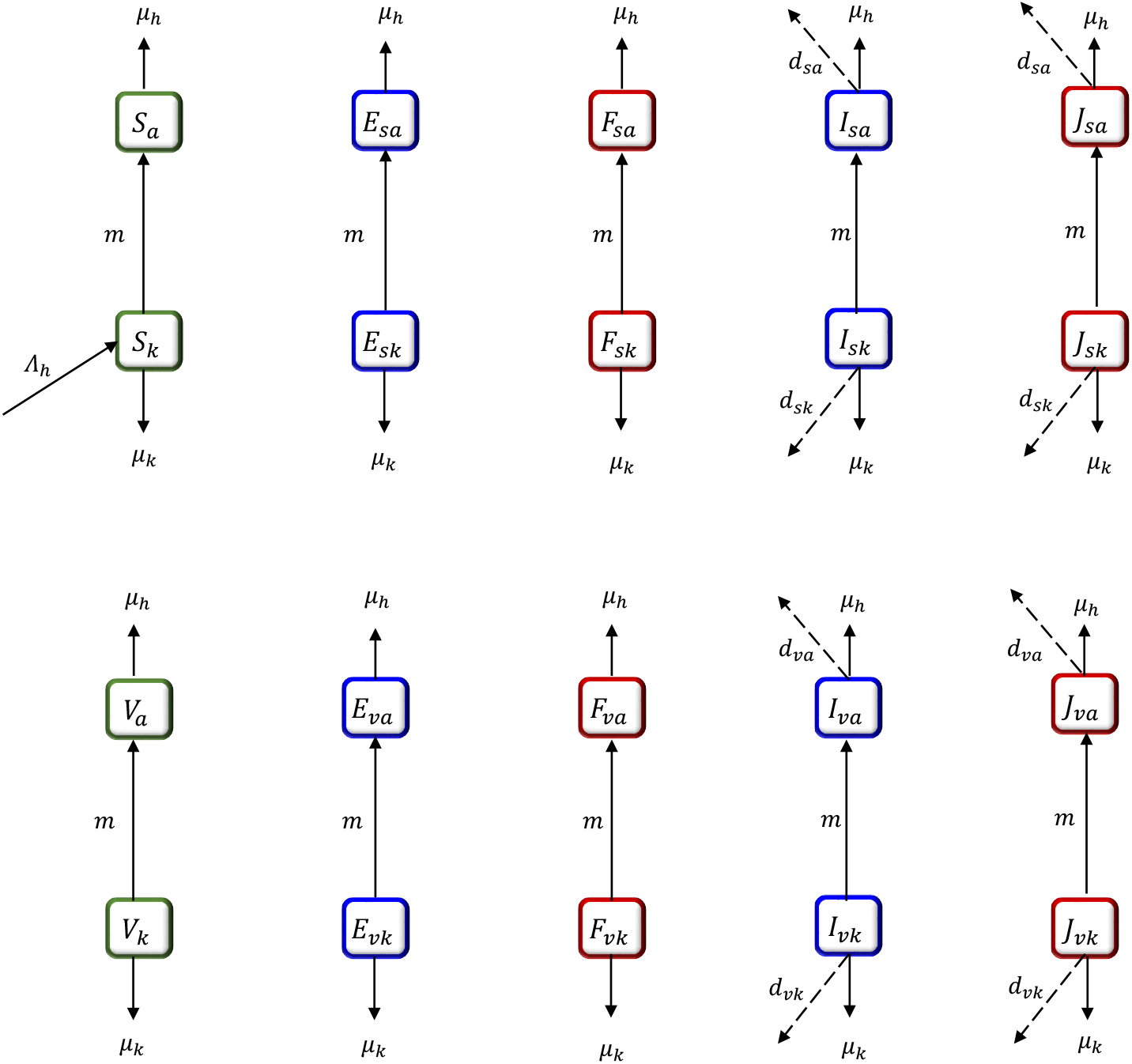
Transfer diagram between the naive-immune juvenile human population and the mature human population. Dashed lines represent disease-induced mortality.

An unvaccinated susceptible mature human (*S*_*a*_) can be exposed to the sensitive malaria strain and transition to (*E*_*sa*_), or be exposed to the resistant strain and transition to (*F*_*sa*_). From (*E*_*sa*_), they can become infectious and transition to (*I*_*sa*_) for the sensitive strain, and from (*F*_*sa*_) to (*J*_*sa*_) for the resistant strain, with an incubation rate of *δ*_*a*_. From the infected state, unvaccinated mature humans may further transition to a treatment class, (*T*_*sa*_), where they receive treatment at rate *a* with anti-malaria drugs with efficacy *P*_*s*_ for the sensitive strain and *P*_*r*_ for the resistant strain. Additionally, infected unvaccinated mature humans may naturally clear their infection at a rate of *σ*_*sa*_, with a proportion *ξ*_*Isa*_ for the sensitive strain and *ξ*_*Jsa*_ for the resistant strain, developing temporary immunity to transition to *R*_*s*_. The remaining fraction, 1 *− ξ*_*Isa*_ for the sensitive strain and 1 *− ξ*_*Jsa*_ for the resistant strain, of infected, unvaccinated, mature humans return to *S*_*a*_ to become susceptible, unvaccinated, mature humans again. Moreover, any recovered mature human will experience a natural loss of immunity at a rate of *ω* and become susceptible again. Vaccinated susceptible mature humans (*V*_*a*_) undergo similar transition states to *S*_*a*_, moving from their corresponding exposed, infected, treatment, to recovered classes.

The equations governing the dynamics of unvaccinated naive humans take the following form

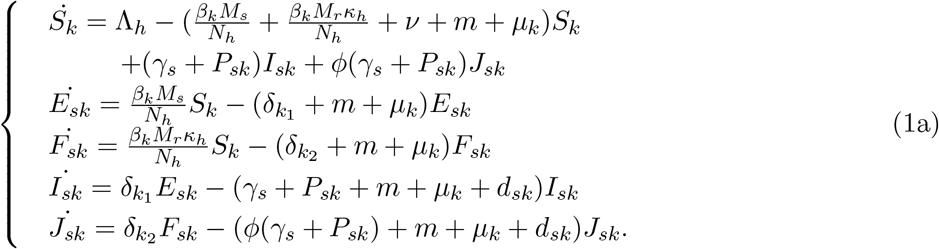

The equations governing the dynamics of unvaccinated mature humans take the following form:

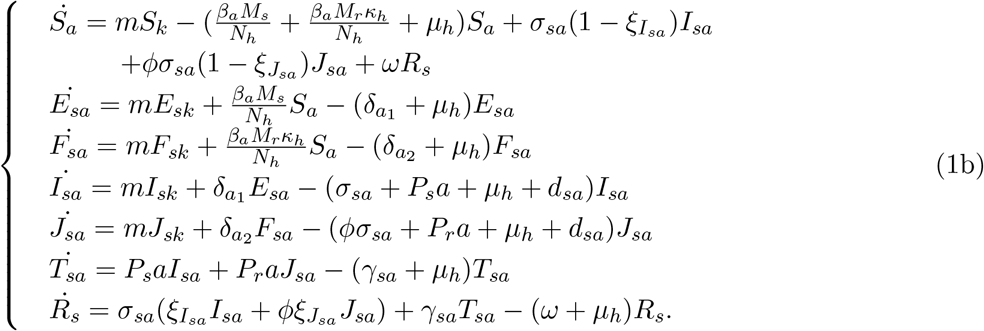

The equations governing the dynamics of vaccinated naive humans take the following form

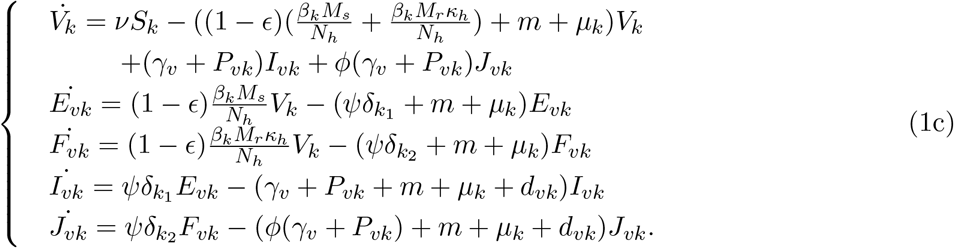

The equations governing the dynamics of vaccinated mature humans take the following form:

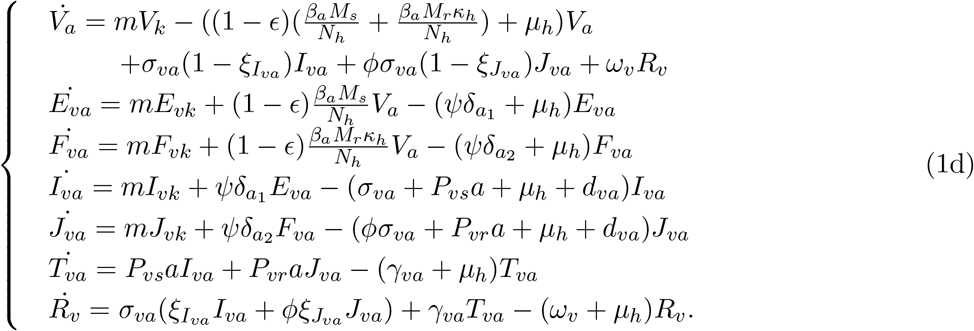

The equations that govern the mosquito dynamics take the following form:

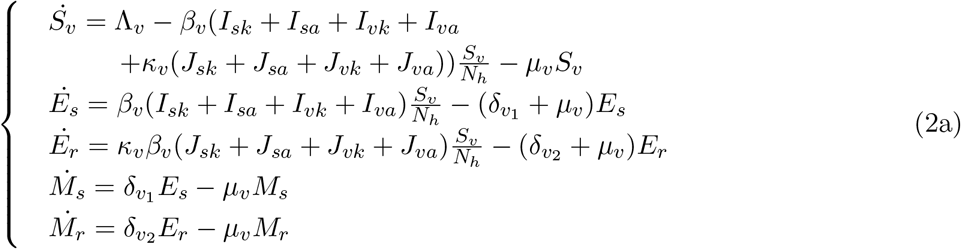

## 3 Model Analysis

In this section, we compute the basic reproduction numbers for the sensitive parasite strain, 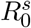, and the resistant parasite strain, 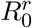, using the next-generation matrix (NGM). Additionally, the invasion reproduction numbers, labeled as 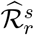 and 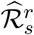, are calculated using the next-generation matrix. In this context, 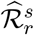 denotes the threshold determining whether the resistant strain can infiltrate the endemic equilibrium limited to only sensitive strains, while 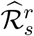 denotes the potential of sensitive strains to infiltrate the endemic equilibrium composed only of resistant strains.

### 3.1 Disease Free Equilibrium

Let

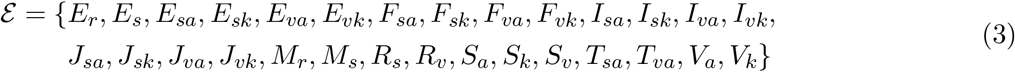

denote an equilibrium of the system. The system has the following disease-free equilibrium (DFE):

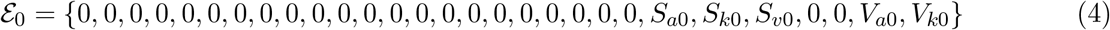

where,

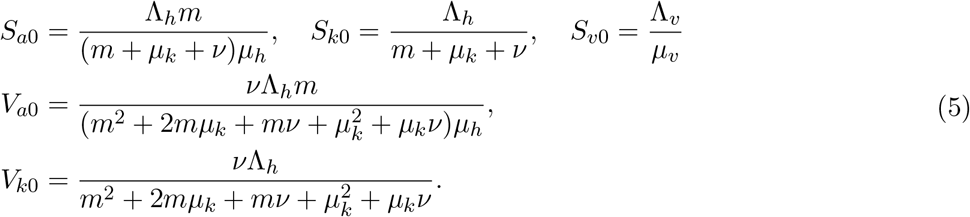

#### Theorem 3.1.

*The disease-free equilibrium Eq. (4) is locally asymptotically stable in the absence of disease*.

*Proof*. First we consider a subsystem of the full model in the absence of disease:

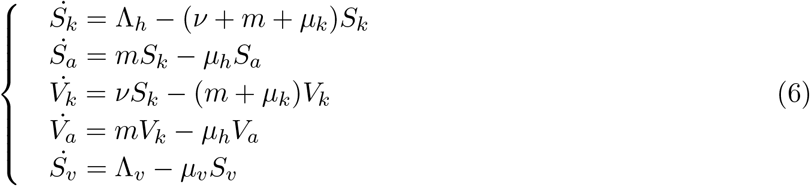

Linearizing Eq. (6) yields the Jacobian matrix,

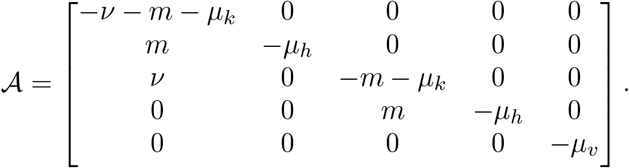

Given that matrix 𝒜 is lower triangular, the eigenvalues are determined directly from the diagonal elements. Since all these diagonal elements are negative, it follows that the disease-free equilibrium Eq. (4) is locally asymptotically stable.

### 3.2 Basic reproduction numbers

The basic reproduction numbers for the sensitive parasite strain, 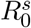, and the resistant parasite strain, 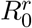, are computed using the next-generation matrix (NGM) [32]. Similarly, the reproduction numbers when interventions such as treatment and vaccination are in place are computed for the sensitive strain,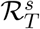, and for the resistant strain 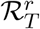.

The intervention reproduction number for the sensitive strain of infection is given by,

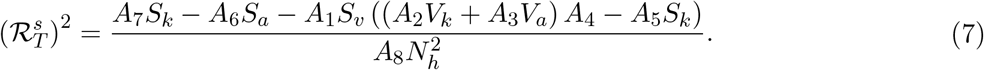

The intervention reproduction number for the resistant strain of infection is given by,

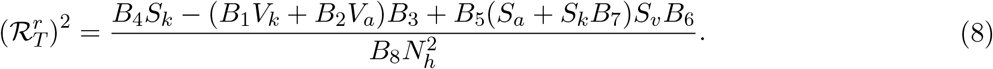

where the durations of infection are represented by the following parameters,

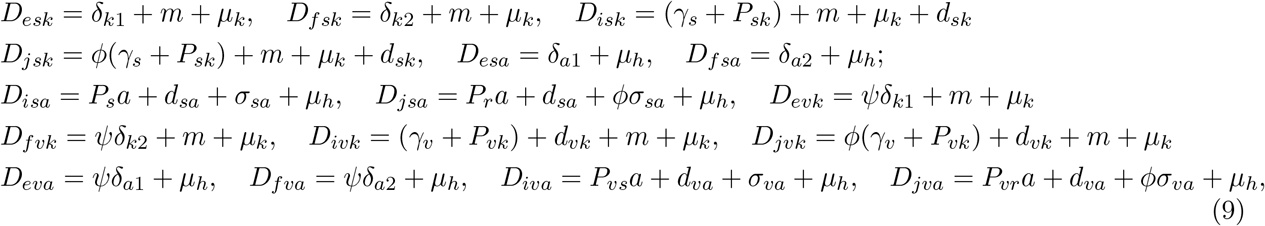

and other parameters are combined and written as,

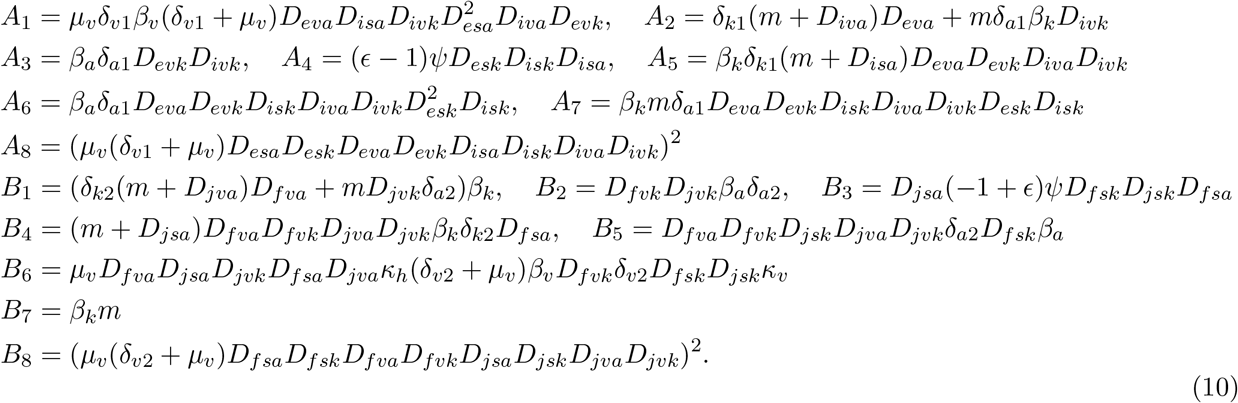

The basic reproduction numbers 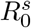 and 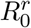 are given using equations (7) and (8) when all intervention parameters, such as *P*_*vs*_ = 0 and *ϵ* = 0 are set to zero.

### 3.3 Invasion reproduction numbers

The basic reproduction number alone is insufficient to determine the competitive outcome between resistant and sensitive strains. In studying models for such cases, a key objective is to identify which infections can invade and persist within a population already harboring other infections. Invasion reproductive numbers (IRNs), which are related to the stability of boundary endemic equilibria, can address this issue. Thus, we derive the IRNs alongside the basic and intervention reproduction numbers. The quantities 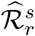 and 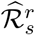 serve as thresholds for determining whether the resistant strain can invade the sensitive-strain boundary equilibrium and vice versa. This derivation utilizes the next-generation approach, substituting the disease-free equilibrium with either the sensitive-only or the resistant-only boundary equilibrium into the NGM.

## 4 Parameterization and Sensitivity Analyses

The descriptions of the model parameters and the sensitivity analyses highlighting the relative significance of these parameters are provided in this section. The majority of the model parameters have been sourced from existing literature. The parameter descriptions and their sources are summarized in Table 2. The efficacy of current antimalarial drugs can vary depending on factors such as the region, primarily due to differing levels of drug resistance. However, in most areas, ACTs and artemisinin derivatives are effective against both drug-sensitive and resistant parasites [27, 33]. ACT drug efficacy rates for children under five in high-transmission areas are consistently high, averaging around 98-99% [13]. The drug treatment efficacies *P*_*sk*_, *P*_*vk*_, *P*_*vr*_, *P*_*s*_, and *P*_*r*_ are defined relative to *P*_*vs*_, assuming that drug treatment is most effective for the human population infected with the sensitive strain, regardless of their vaccination status. If the malaria strain is not fully responsive to the drug, *a · P*_*j*_ quantifies the rate at which the infection clears with treatment, where *j* = *vs, sk, vk, vr, s, r*. Here, a value of *P*_*j*_ close to 1 indicates that the strain is sensitive to the drug, while a value closer to 0 suggests the development of resistance to the treatment. After being exposed to an infective bite by a female Anopheles mosquito, symptoms typically emerge within 7 to 30 days, with shorter incubation periods for *P. falciparum* and longer for *P. falciparum* [4]. We incorporate the reduction factors *κ*_*h*_, *κ*_*v*_, and *ϕ* to clearly distinguish between the transmission and recovery dynamics of resistant and sensitive strains. The parameters *κ*_*h*_ and *κ*_*v*_ specifically reduce the transmission rates for humans and mosquitoes, respectively, in the resistant strain compared to the sensitive strain. Meanwhile, *ϕ* further differentiates the strains by decreasing the natural recovery rate for the resistant strain relative to the sensitive strain. Among the assumed parameters values, *ν* was careful determined to simulate a realistic vaccination rate Fig. B.1 (Appendix B).

The accuracy of results from mathematical and computer models of biological systems is often compromised by uncertainties in experimental data, and traditional single-parameter or local sensitivity analyses fail to address this comprehensively. Global sensitivity analysis techniques, however, allow for the identification and control of these uncertainties in a multi-dimensional parameter space. Latin Hypercube Sampling (LHS), as introduced in [16], is a stratified Monte Carlo sampling technique that ensures each parameter interval is sampled exactly once across multiple simulations without replacement. This method allows for an unbiased estimate of the average model output with a limited number of samples. When combined with the Partial Rank Correlation Coefficient (PRCC) technique [15], the LHS method provides an unbiased estimate of the average model output while sampling every parameter. Using LHS to sample the parameters in Table 2, we apply PRCC to conduct a global sensitivity analysis of the reproduction numbers and the proportion of the population infected with both sensitive and resistant strains. We use PRCC since it is an appropriate statistical measure in sensitivity analyses when parameters exhibit a nonlinear and monotonic relationship with the output measure, as illustrated in Appendix A (Fig. A.1).

Fig. 4 shows the resulting PRCC values for the reproduction numbers (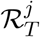, *j* = *s, r*) and proportion of the population infected with both sensitive and resistant strains. In this context, parameters with higher-magnitude PRCC values are generally more influential than those with lower PRCC values. A positive PRCC value indicates a direct relationship with the output measure, meaning that an increase in the parameter is likely to increase the respective output measure. Conversely, a negative PRCC value reflects an inverse relationship, where an increase in the parameter is likely to decrease these output measures. By performing a z-test on the PRCC values, we verify that higher magnitude PRCC values generally indicate a stronger impact on the output measures.

**Figure 4:**
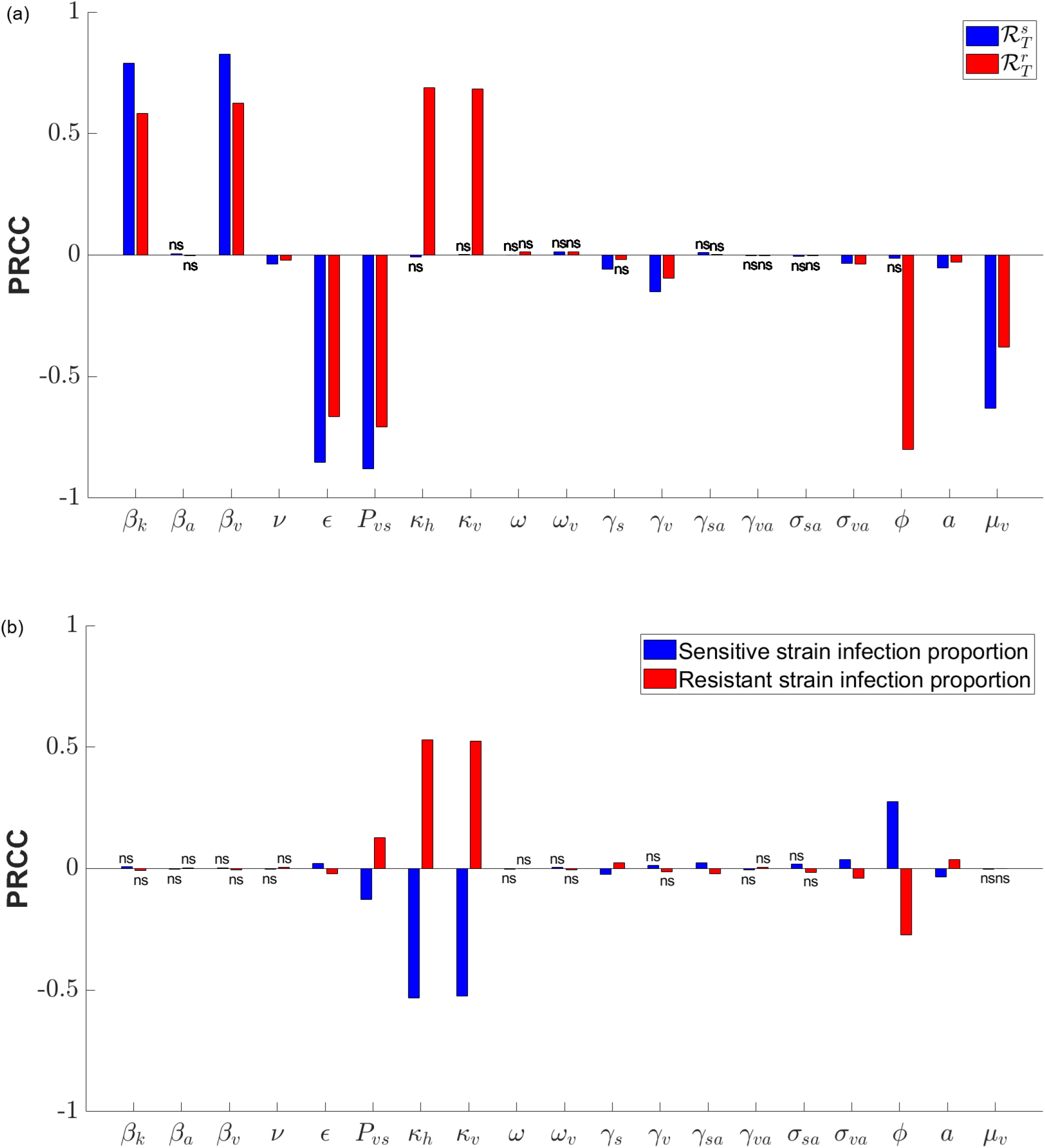
Sensitivity analysis of the model using Partial Rank Correlation Coefficient (PRCC) values for each parameter in the Latin Hypercube Sampling with 10,000 samples. (a) PRCC values corresponding to reproduction numbers 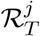, *j* = *s, r*. (b) PRCC values corresponding to the proportion of the population infected with both sensitive and resistant strains after 100 years. PRCC values marked as “ns” are not significant (*p*-test value *≥* 0.05). Each parameter is represented by a pair of bars, with sensitive strains shown in blue and resistant strains in red.

The PRCC results indicate that the most influential parameters for both 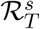 and 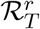 are the transmission rates from mosquitoes to naive humans (*β*_*k*_) and from humans to mosquitoes (*β*_*v*_), the vaccination rate (*ν*) and efficacy (*ϵ*), the effectiveness of drug treatments (*P*_*vs*_), recovery rates (*γ*), the average time required to clear the sensitive strain (*a*), and the mosquito death rate (*μ*_*v*_). Additionally, the reduction factors in transmission rates (*κ*) and natural recovery rates for resistant strain compared to sensitive strain (*ϕ*) have the most significant impact on 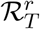, which are not influential for 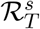. When examining the PRCC plot for the proportion of the population infected with both sensitive and resistant strains, we observe that only drug efficacies (*P*_*vs*_), along with the reduction factors in transmission rates (*κ*) and natural recovery rates for resistant strains (*ϕ*), and the average time required to clear the sensitive strain (*a*), significantly impact the PRCC values.

## 5 Numerical Explorations

In this section, we present numerical simulations to illustrate the dynamic behavior of our model. The parameters used in these simulations are listed in Table. 2. We investigate how changes in transmission rates under different treatment and vaccination efficacies affect the basic reproduction numbers, as well as the intervention and invasion reproduction numbers. Additionally, we explore the effect of changing treatment and vaccination efficacies on the reproduction numbers and the fraction of the population infected with either strain.

In the following Fig. 5, we present the numerical solution of the model, illustrating the exposed and infected populations for both sensitive and resistant strains over a period of 5 years. The infectious population for both strains reaches a peak at approximately 7 to 8 months. Although the infectious population of the sensitive strain initially rises across all categories—vaccinated and unvaccinated, naive and mature individuals, it eventually declines and dies out. In contrast, after an initial decline following the peak, the resistant strain’s infectious population continuously persists across all four categories. Over time, the resistant strain surpasses the sensitive strain, ultimately prevailing and not dying out. In Fig. 6 (b), this transition, where the fraction of the infected population with the resistant strain surpasses that with the sensitive strain, is noticed. Additionally, this shift in the mosquito population follows a similar pattern but occurs at a later time (Fig. 6 (d)).

**Figure 5:**
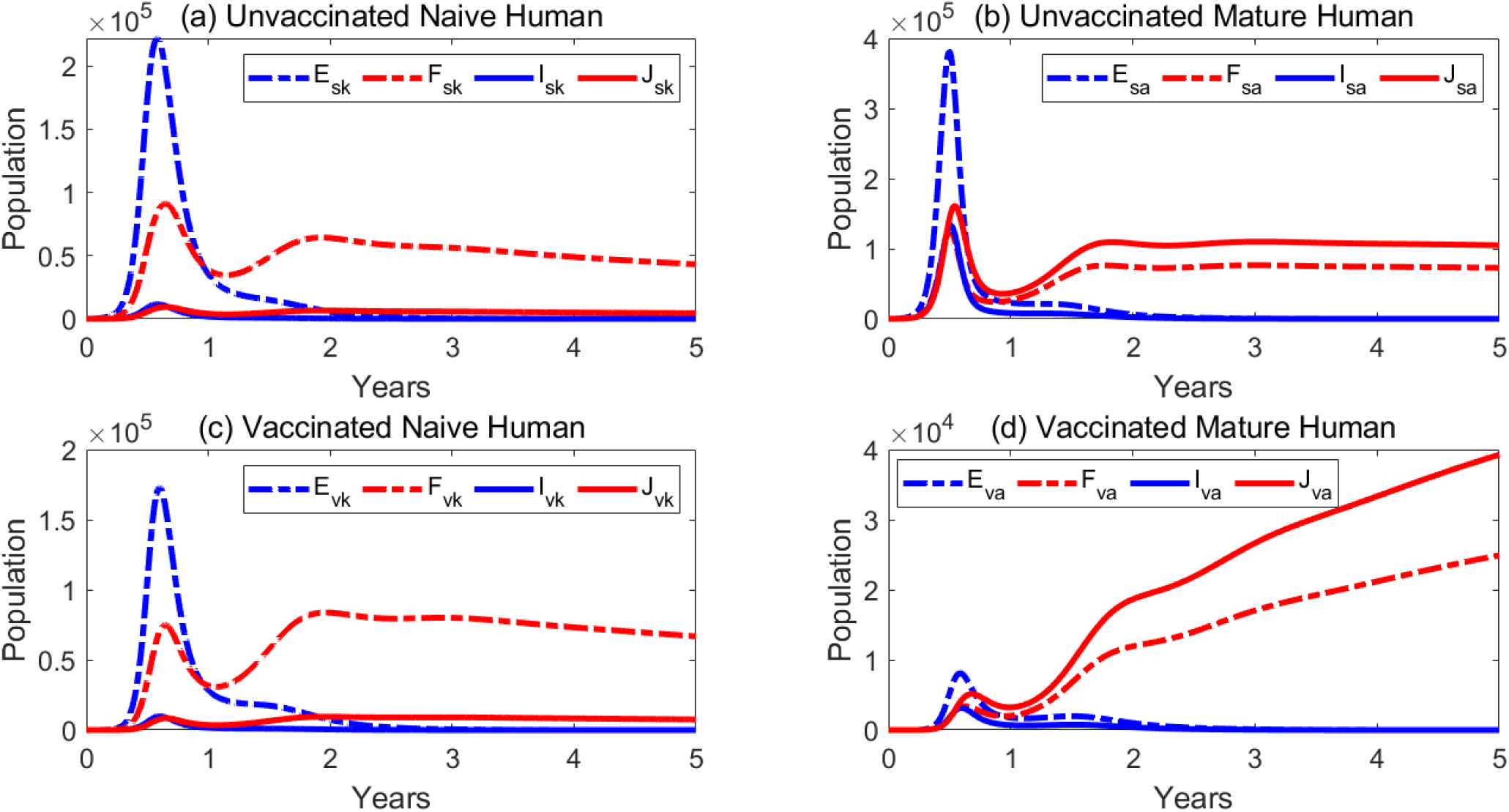
Numerical solutions of the model of the exposed and infected populations for both sensitive and resistant strains over a period of 5 years. Resistant strains are in red and sensitive strains in blue, with dashed lines for exposed populations and solid lines for infected populations. All parameters are maintained at baseline values as listed in the Table 2. We consider an initial human population of *N*_*h*_(0) = 5 million and a mosquito population of *N*_*v*_(0) = 15 million. Additional initial conditions are as follows: *S*_*k*_(0) = 0.40 *× N*_*h*_(0), *S*_*a*_(0) = 0.60 *× N*_*h*_(0), *S*_*v*_(0) = 3 *× N*_*h*_(0); *E*_*s*_(0) = 50; *E*_*r*_(0) = 50; *M*_*s*_(0) = 100; *M*_*r*_(0) = 100. All other classes are initialized to zero.

**Figure 6:**
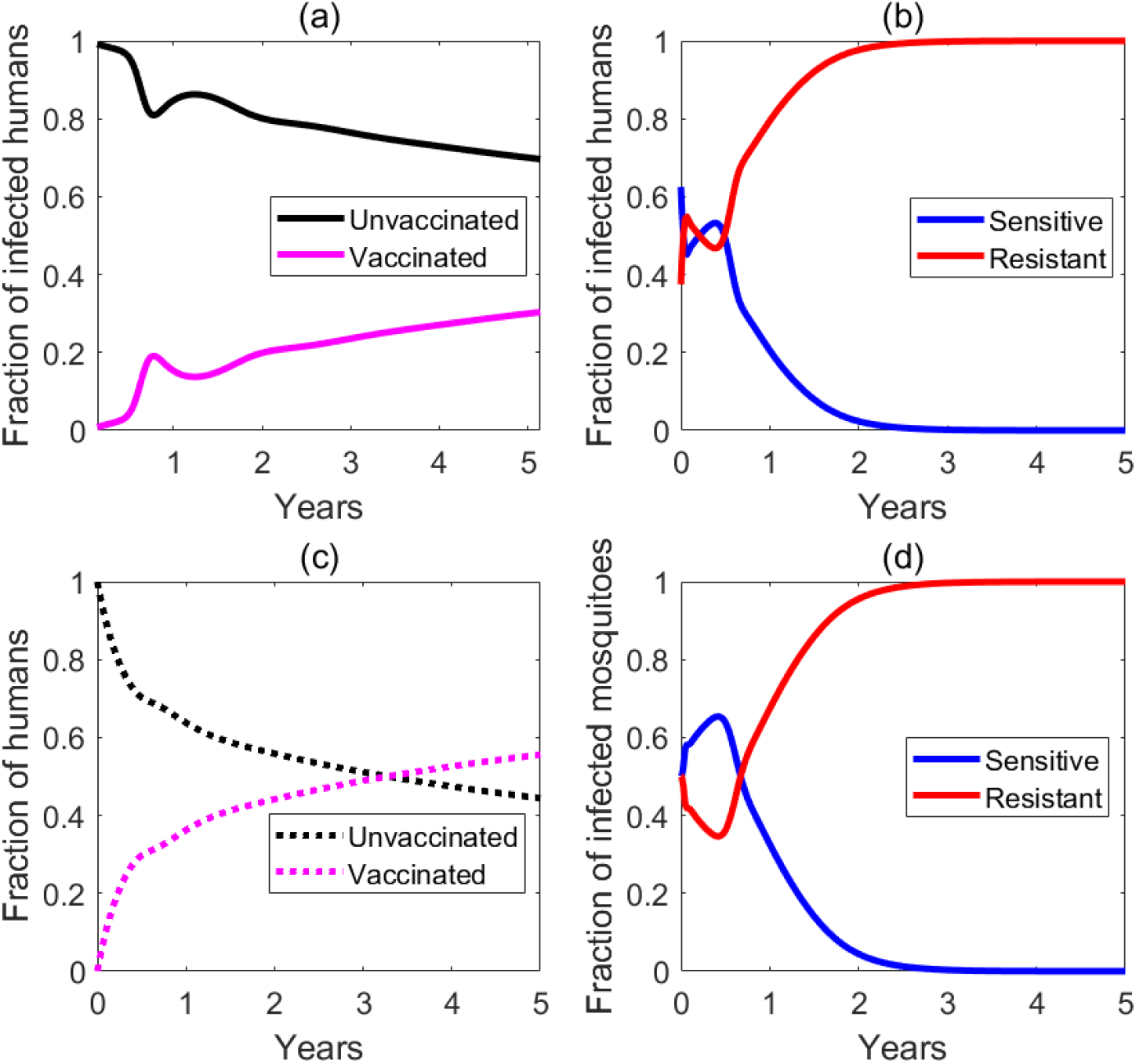
Fraction of the population over 5 years. (a) Represents the fraction of the infected human population categorized by vaccination status. (b) Shows the fraction of the infected human population that is infected with resistant and sensitive strains. (c) Illustrates the changing ratio of vaccinated and unvaccinated individuals within the overall human population over time. (d) Represents the fraction of the infected mosquito population that is infected with resistant and sensitive strains. All parameters are maintained at baseline values as listed in the Table 2.

In our simulations, the vaccination rate *ν* has been carefully chosen to reflect realistic vaccination expectations in endemic countries. As parameterized, our model demonstrates that approximately 86.54% of the population will be vaccinated after 100 years (Fig. B.1). Moreover, Figures Fig. 6 (a) and Fig. 6 (c) reveal that as the proportion of vaccinated individuals increases over time, the fraction of the infected population that is vaccinated initially rises due to the growing number of vaccinated children. However, this eventually reaches a steady state, where the proportion of the vaccinated population remains significantly lower than that of the unvaccinated population.

In the next set of numerical examples, we examine the impact of changing parameters on the reproduction numbers for the sensitive parasite strain, 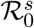, and the resistant parasite strain, 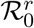. In the absence of any interventions, such as treatment (*P*_*vs*_ = 0) or vaccination (*ϵ* = 0), 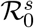 is higher than 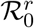 for any given level of transmission (Fig. 7). As expected, when the transmission rate between humans and mosquitoes increases, the basic reproduction numbers also increase. However, with the implementation of treatment and vaccination, the trend reverses. 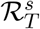 becomes lower than 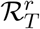 for the same transmission level, and the reproduction numbers for both strains are significantly reduced, demonstrating the effectiveness of these interventions (Fig. 8). Furthermore effects of treatment efficacy *P*_*vs*_ and vaccination efficacy *ϵ* can be significant on both 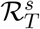 and 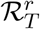 (Fig. 9). Under low vaccine efficacy the sensitive strain is more responsive to drug treatments compared to the resistant strain (Fig. 9 (b)). However, when drug efficacies are low, very high vaccine efficacies are more effective against the resistant strain (Fig. 9 (a)).

**Figure 7:**
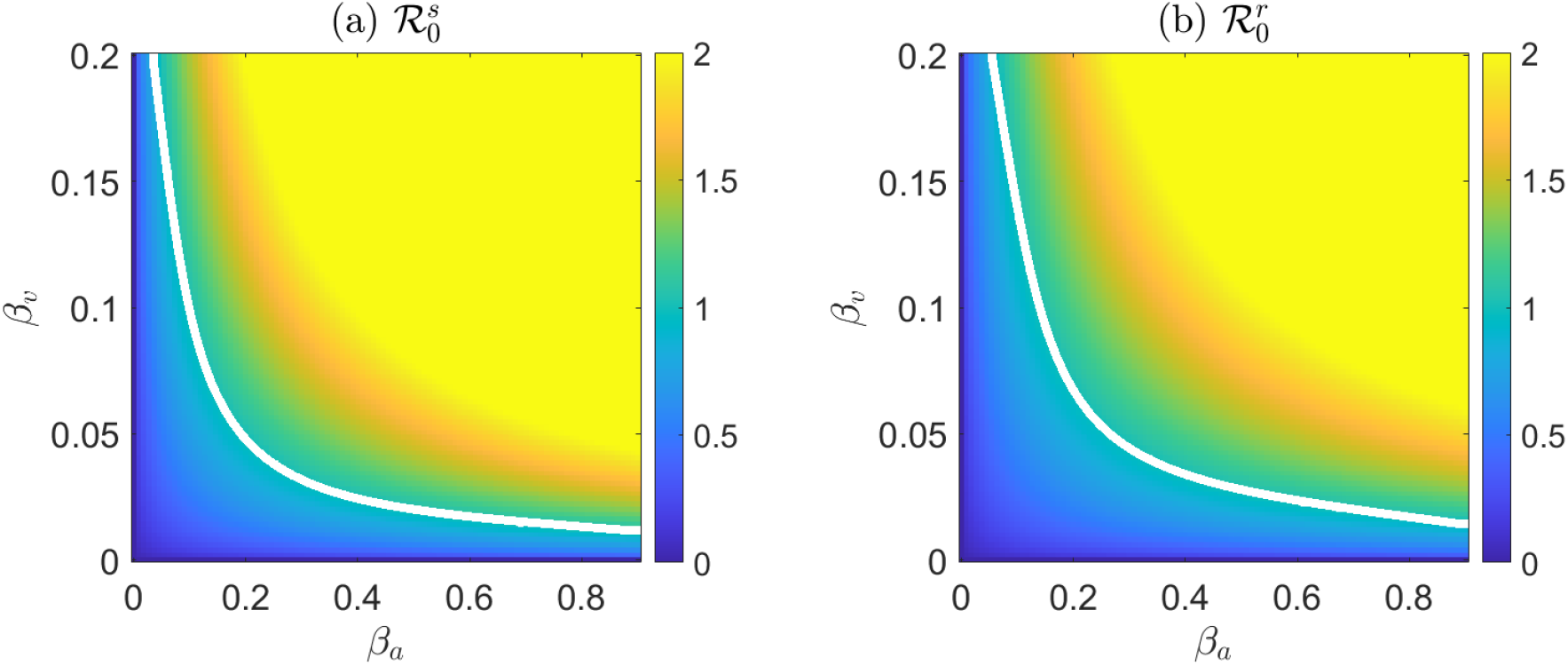
Heatmap illustrating the impact of variations in transmission rates for humans (*β*_*a*_ and *β*_*k*_) and mosquitoes (*β*_*v*_) on the basic reproduction numbers for both sensitive 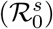 and resistant strains 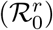 in the absence of treatment and vaccination, i.e., *P*_*vs*_ = 0 and *ϵ* = 0. Here, *β*_*a*_ is set equal to the transmission rate for humans, while *β*_*k*_ is set at 90% of the human transmission rate. All other parameters are maintained at their baseline values as listed in Table 2. The white curve indicates when *R*_0_ equals 1.

**Figure 8:**
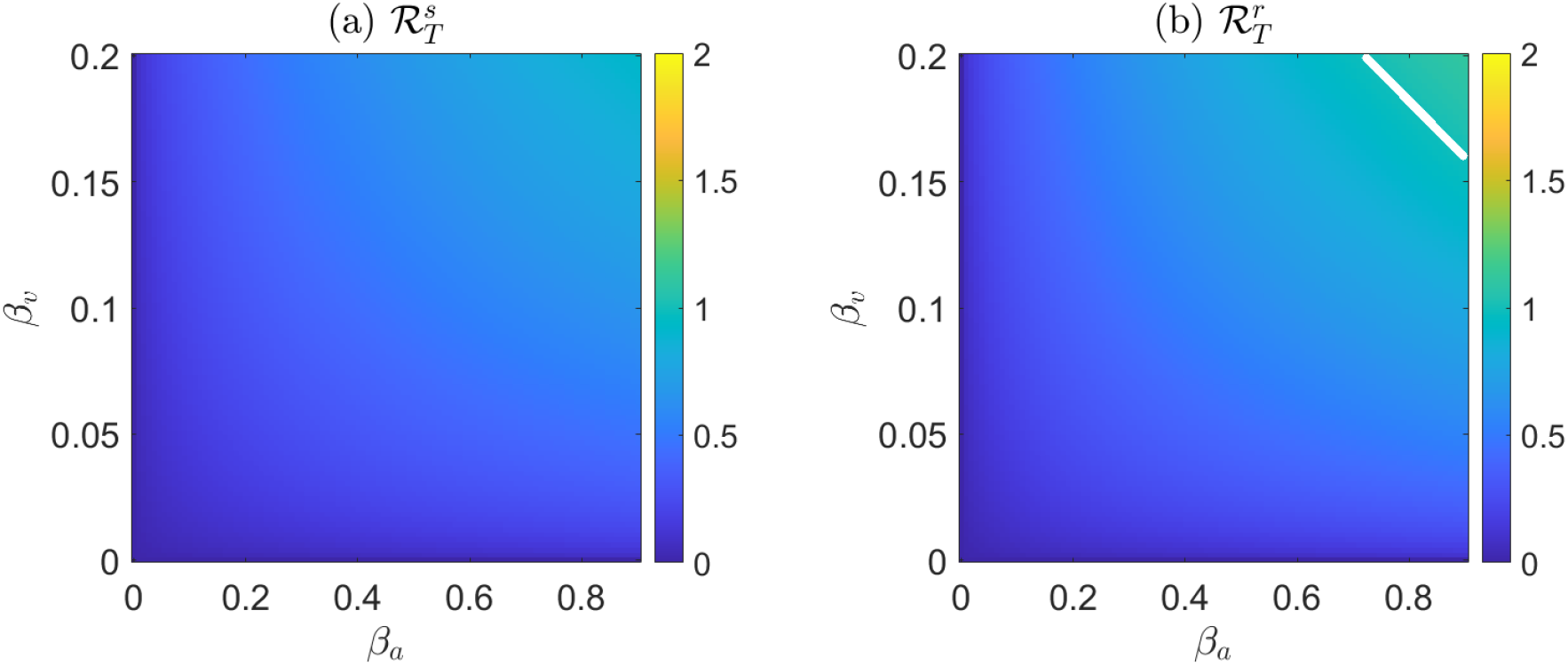
Heatmap illustrating the impact of variations in transmission rates for humans (*β*_*a*_ and *β*_*k*_) and mosquitoes (*β*_*v*_) on the reproduction numbers for both sensitive 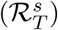 and resistant strains 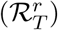 in the presence of treatment and vaccination, where *P*_*vs*_ = 1 and *ϵ* = 0.75. Here, *β*_*a*_ set equal the transmission rate for humans, while *β*_*v*_ is set at 90% of the human transmission rate. All other parameters are maintained at their baseline values as listed in Table 2. The white curve indicates when *R*_*T*_ equals 1.

**Figure 9:**
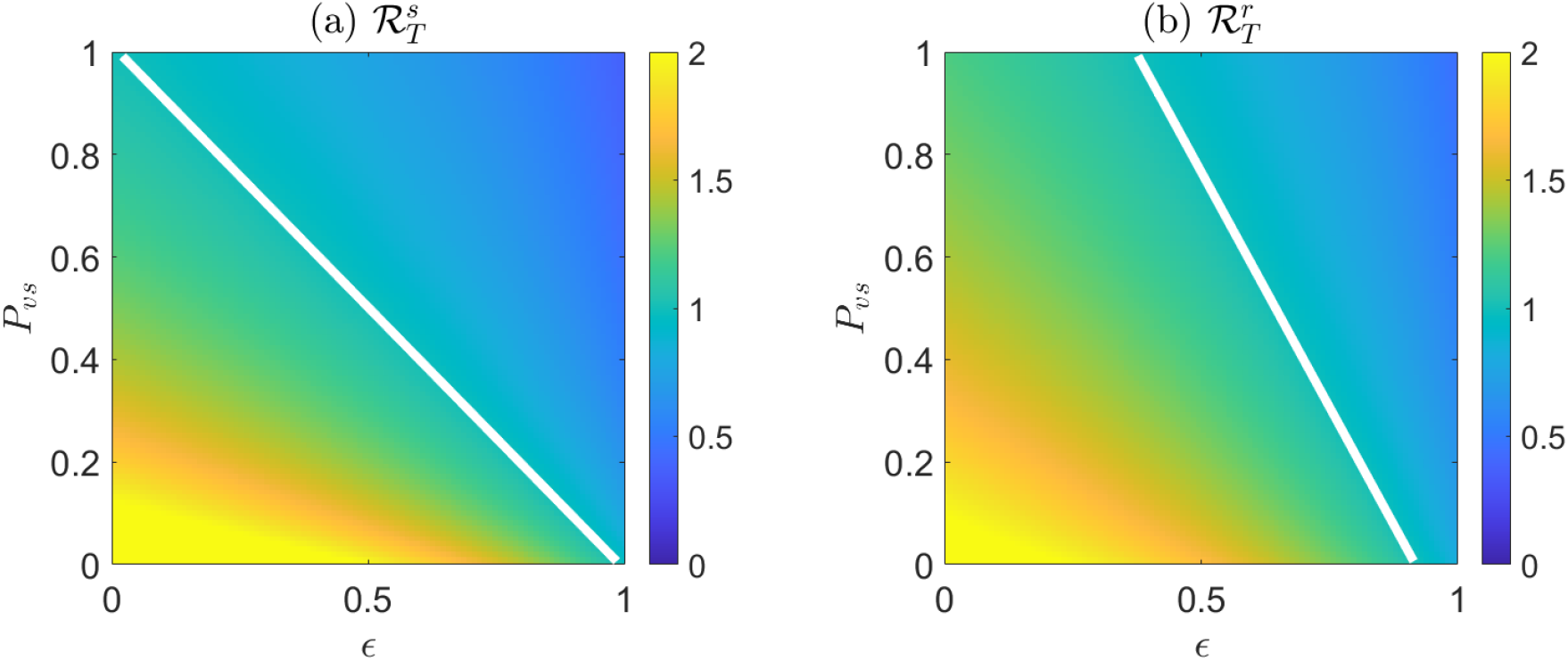
Heatmap illustrating the impact of variations in vaccination efficacy (*ϵ*) and treatment efficacy (*P*_*vs*_) on the reproduction numbers for both sensitive 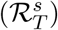 and resistant strains 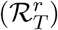. All other parameters are maintained at their baseline values as listed in Table 2. The white curve indicates when *R*_*T*_ equals 1.

We observe a switch between the prevalence of sensitive and resistant infections after a certain treatment efficacy threshold, around 20%, is reached. This trend is consistent across different vaccination efficacies (Fig. 10). The switch between sensitive and resistant infections at a certain treatment efficacy threshold highlights the complex interplay between drug efficacy and resistance evolution. While highly effective treatments are crucial for eliminating sensitive infections, they can inadvertently contribute to the rise of resistance. Conversely, higher vaccination efficacy results in a lower percentage of the population becoming infected with either strain, with no surge of the resistant strain if the vaccination efficacy exceeds 90% ((Fig. 10 (c)). These findings underscore the importance of a synergistic approach combining both treatment and vaccination is the most effective strategy for mitigating malaria in a population.

**Figure 10:**
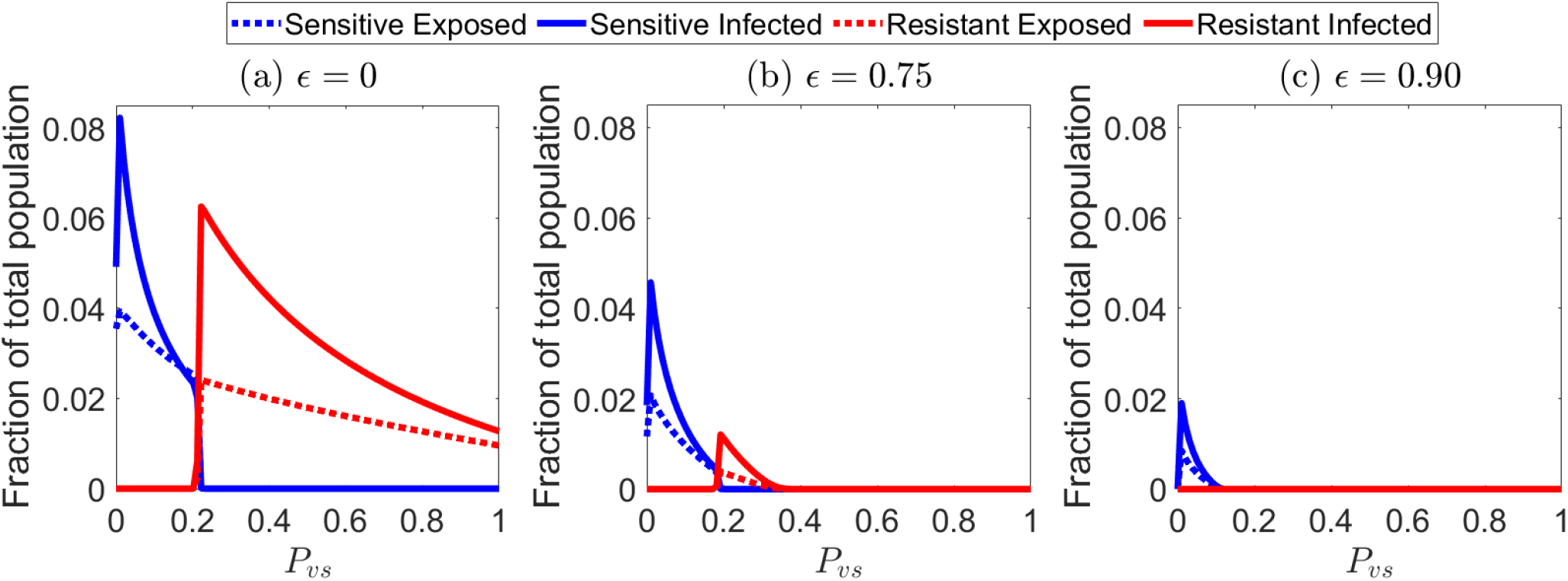
The effect of varying treatment efficacy on the fraction of the population infected with sensitive and resistant strains is examined over a period of *t* = 100 years, during which both treatment and vaccination have been implemented in the system. Three scenarios are considered, each representing different levels of vaccination efficacy: 0, 0.75, and 0.90. All other parameters are maintained at their baseline values from Table: 2

In Fig. 11, we further explore the fraction of the infected population as *P*_*vs*_ and *ϵ* change. We observe that starting with an initial condition of only the sensitive strain, only the sensitive strain exists and eventually dies out when *P*_*vs*_ and *ϵ* are high enough (Fig. 11, (a)). Similarly, if there is an initial condition of only the resistant strain, then only the resistant strain exists before eventually dying out at much higher levels of drug efficacy but lower vaccine efficacy (Fig. 11, (b)). When both strains are initially present, the sensitive strain outcompetes the resistant strain until drug efficacy surpasses 0.20 (Fig. 11, (c)), which is consistent with observations in Fig. 10 and Fig. 12.

**Figure 11:**
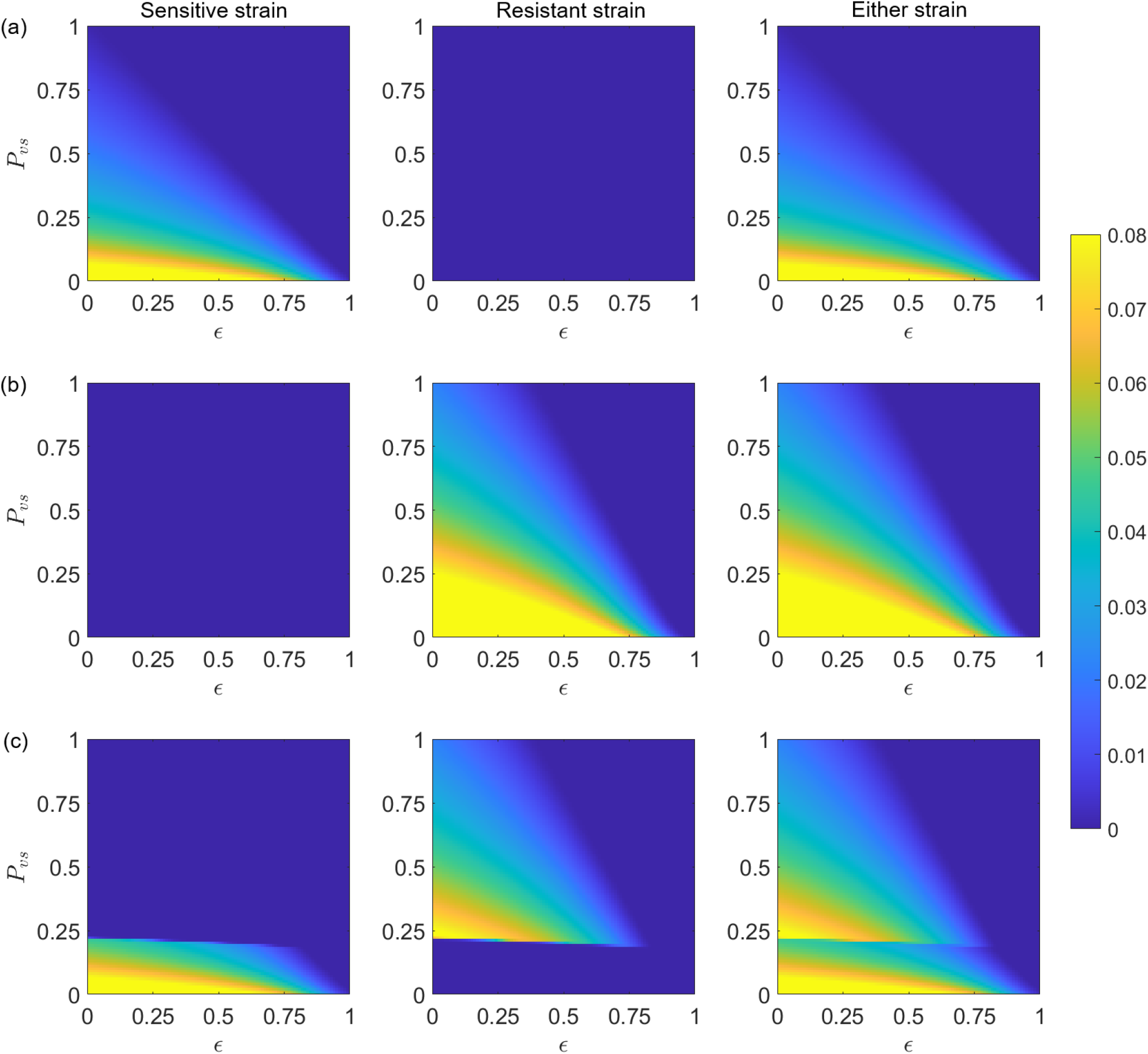
Fraction of the population infected with the sensitive strain (first column), the resistant strain (second column), and either strain (third column) after a period of *t* = 100 years. Initial conditions are set as follows: *S*_*k*_(0) = 0.40 *× N*_*h*_(0), *S*_*a*_(0) = 0.60 *× N*_*h*_(0), *S*_*v*_(0) = 3 *× N*_*h*_(0); For (a) *E*_*s*_(0) = 50, *M*_*s*_(0) = 100, with all other classes are initialized to zero. For (b) *E*_*r*_(0) = 50, *M*_*r*_(0) = 100, with all other classes are initialized to zero. For (c) *E*_*s*_(0) = 50, *E*_*r*_(0) = 50, *M*_*s*_(0) = 100, *M*_*r*_(0) = 100, with all other classes are initialized to zero. All other parameters are maintained at their baseline values from Table 2.

**Figure 12:**
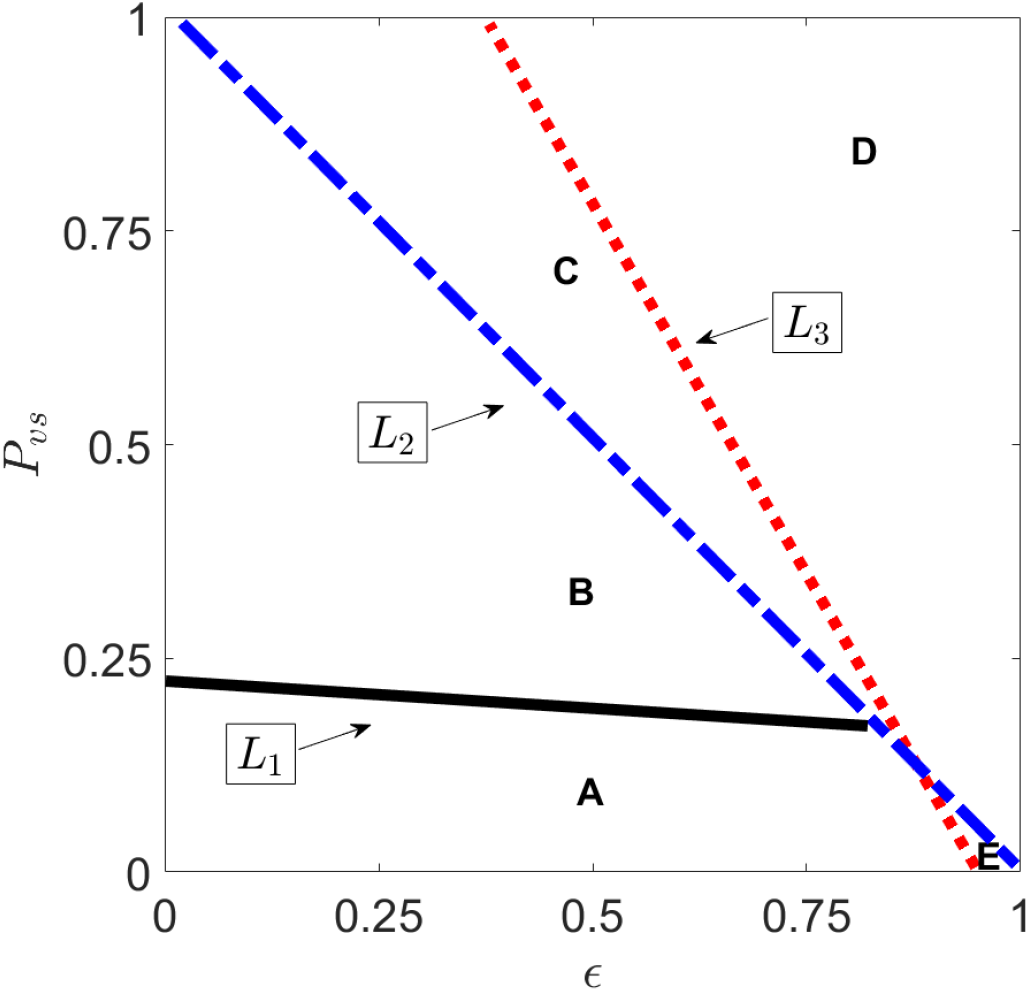
Malaria strain competitive outcomes across varying vaccination (*ϵ*) and treatment efficacies (*P*_*vs*_) over a period of 100 years. All other parameters are kept the same as in Table: 2. Threshold lines dividing the parameter space into different regions are 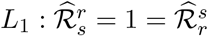 in black, 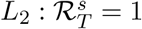 in dashed blue, and 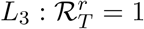 in dotted red. In regions **A** and **E**, the sensitive strain outcompetes the resistant strain. If the resistant strain is not introduced, the sensitive strain will persist. In regions **B** and **C**, the resistant strain outcompetes the sensitive strain. The disease dies out in region **C** if the resistant strain is not introduced, in region **E** if the sensitive strain is not introduced, and in region **D** regardless of the strain introduced.

In Fig. 12, we illustrate the competitive outcomes of malaria strains under varying vaccination (*ϵ*) and treatment efficacies (*P*_*vs*_), taking into account the intervention and invasion reproduction numbers for both the sensitive and resistant strains. Threshold lines, defined by reproduction numbers, divide the parameter space into different regions. In region **A**, the sensitive strain outcompetes the resistant strain. If the resistant strain is not introduced, the sensitive strain will persist. Here, both the reproduction numbers for the sensitive and the resistant strains, denoted as 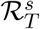 and 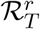 respectively, are greater than 1. However, the invasion reproduction number for the sensitive strain invading the resistant-only equilibrium, denoted as 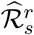, is also greater than 1, while the invasion reproduction number for the resistant strain invading the sensitive-only equilibrium, denoted as 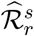, is less than 1. The line *L*_1_ indicates where the switch in the invasion reproduction numbers takes place. At *L*_1_, 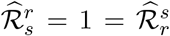. In region **B**, the resistant strain outcompetes the sensitive strain. If the resistant strain is not introduced, the sensitive strain will persist. Here, 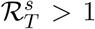, 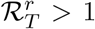, and 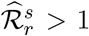 1 but 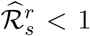 1. The threshold 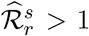 indicates that the resistant strain is able to invade the sensitive-strain boundary equilibrium. In region **C**, 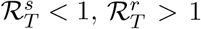, and 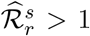 but 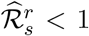. The dashed line *L*_2_ indicates where the switch in the reproduction number for the sensitive strain takes place; 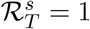. If the resistant strain is not introduced, the disease will die out, resulting in no infection with either strain. In region **D**, both 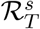 and 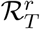 are less than 1, indicating that the disease will die out regardless of the introduction of either strain. Finally, region **E** represents a small portion of the parameter space where 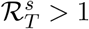 but 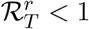. In region **E**, if the sensitive strain is not introduced, the disease will die out, leading to no infections with either strain. However, if the resistant strain is introduced, the disease will persist.

## 6 Conclusion

Malaria continues to be a burden in many parts of the world. Following our preceding discussion and the recent World Malaria Report [35], which shows an increase in malaria cases and deaths worldwide over the past reporting period, it is clear that reducing infection and disease burden in infants and children, the groups bearing the highest burden of the disease, is increasingly urgent. The recent groundbreaking RTS,S malaria vaccine (trade name Mosquirix) and the latest advancement in malaria vaccination, the R21/Matrix-M™ vaccine, which have demonstrated impressive efficacy, show promise. On the treatment front, existing antimalarial drugs like Dihydroartemisinin (DHA), artesunate, and artemether—derivatives of artemisinin—remain critical in managing and treating the disease with concerns about drug resistance which could undermine the effectiveness of these treatments.

The primary objective of this research is to offer new insights into how malaria vaccination coverage influences disease prevalence and transmission dynamics. We found that our extended SEIR model is mostly impacted by transmission rates, vaccination rate and efficacy, the effectiveness of drug treatments, scale factors that reduce transmission and natural recovery rates for the resistant strain, and mosquito death rates.(Fig. 4). Hence, intervention strategies should focus on increasing vaccination coverage, enhancing drug efficacy, addressing factors contributing to resistant strains, and implementing effective vector control measures, e.g.,[18].

The numerical simulations of the model reveal complex interplay between drug efficacy and the evolution of resistance (Figs. 9, 10, 11, and 12). In particular, Fig. 10 shows increased drug efficacy leads to the emergence of the resistant strain, whereas higher vaccine efficacy reduces the proportion of the population infected with either strain and, at sufficiently high levels, can prevent the emergence of resistant strains entirely (Fig. 10 (c)). These findings are further supported by the plots of the basic and invasion reproduction numbers, which confirm the observed trends (Figs. 9 and 12). The model observations underscore the importance of nuanced approaches to disease management that prioritizes both treatment efficacy and resistance prevention. Furthermore, the findings suggest that combining moderate drug efficacy with other strategies, such as vaccination, transmission reduction, or integrated treatment plans, could be more effective in controlling both sensitive and resistant infections over the long term.

While our model considers both drug-sensitive and drug-resistant strains in an extended SEIR age-structured framework with malaria vaccination for children, it does not consider vaccination of mature humans. Vaccination strategies targeting across multiple age classes might have significant impacts. It is also worth noting that a big challenge arises from the ease with which Plasmodium can be transported from regions where it is endemic to regions where it is not, due to increased human mobility in the modern world [26]. Future extension of the model considering human mobility will likely lead to additional insights. Temperature and rainfall are well-established factors that significantly impact mosquito population dynamics and the development rate of the malaria parasite within mosquitoes, thereby influencing malaria transmission. Incorporating regional temperature data into the model, especially with seasonal variations e.g., [21], also presents an exciting prospect for future research.

## Data Availability

No new data were generated or analyzed in this study. The parameters used in the model are sourced from existing literature, as cited in the manuscript.

## CRediT authorship contribution statement

**Mahmudul Bari Hridoy:** Conceptualization, Formal analysis, Methodology, Software, Writing – original draft, Writing – review & editing. **Angela Peace:** Conceptualization, Formal analysis, Supervision, Writing – original draft, Writing – review & editing.

## A Parameter sensitivity monotonicity plots

**Figure A.1:**
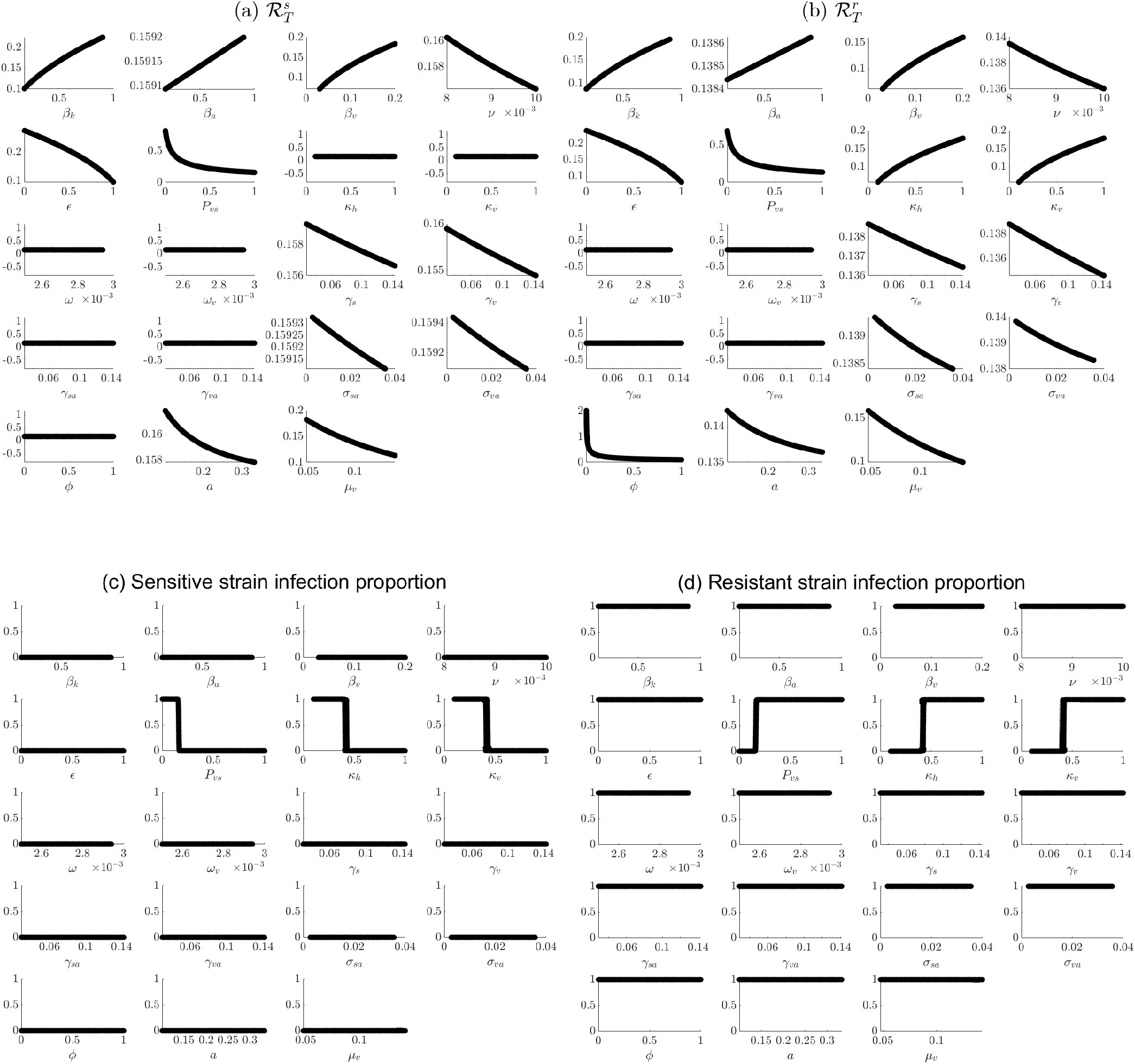
Simulations results showcase that each parameter has a monotonic relationship with the given output measures used in the LHS/PRCC parameter sensitivity analyses. Here, individual parameters were varied, samples using the LHS procedure while all other parameters were fixed at baseline values.

## B Vaccination rate parameterization

**Figure B1:**
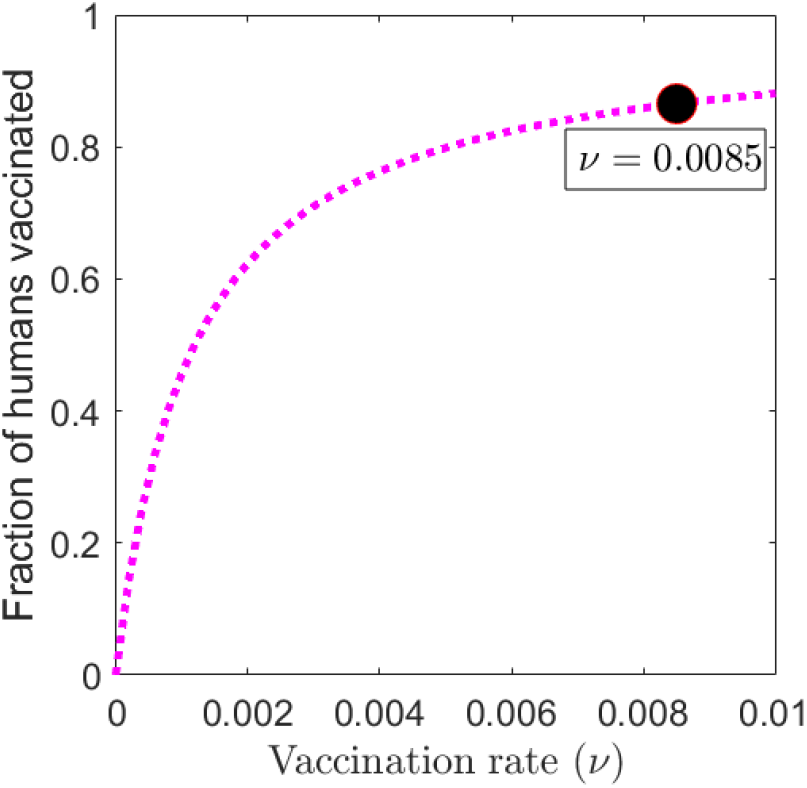
The fraction of the population vaccinated over a 100-year period is graphed as a function of the vaccination rate (*ν*). The black circle represents the baseline vaccination rate (*ν* = 0.0085) used in our simulations, resulting in approximately 86.54% of the population being vaccinated after 100 years. All other parameters are kept at baseline values as listed in Table 2, with initial conditions set as in Figure 5.

